# Handgrip strength as a screening tool for diabetes in resource-constrained settings: a potential solution to overcome barriers to diagnosis

**DOI:** 10.1101/2023.10.19.23297260

**Authors:** Lekan Sheriff Ojulari, Swabirah Enimire Sulaiman, Taofeek Olanrewaju Ayinde, Eniola Riskat Kadir

**Author notes:** Corresponding author LS.

## Abstract

**Background Information:** Diabetes mellitus is an escalating global health concern, especially in low and middle-income countries. The high cost and inaccessibility of diagnostic tools in resource-constrained settings have heightened the need for alternative screening methods. Handgrip strength (HGS), a measure of muscle strength, emerges as a potential non-invasive and affordable screening tool for diabetes, particularly in areas with limited healthcare access.

**Objective:** To investigate the relationship between handgrip strength and blood glucose regulation in non-diabetic young adults and to provide valuable insights into the potential of handgrip strength as a preventive and affordable approach to managing diabetes.

**Methods:** A cross-sectional study was conducted involving 59 students (aged 18-21) from the University of Ilorin, Nigeria. Handgrip strength was measured using a dynamometer, and its relationship with blood glucose regulatory markers, such as fasting blood glucose, 2-hour post-prandial glucose, and HbA1c, was analyzed. Multiple regression models were utilized to examine the potential associations.

**Results:** Findings revealed significant associations between HGS and glucose regulation markers, particularly FBS, among males. In females, the relationship was evident only after adjusting for body mass index (BMI). Furthermore, a notable relationship between HGS and 2-hour post-prandial glucose levels was observed in females but not in males. However, no significant associations were found between HGS and serum insulin levels across genders.

**Conclusion:** Our study introduces HGS as a practical and cost-effective screening tool for blood glucose regulation disorders, aligning with existing literature and offering a personalized approach to management. In resource-constrained settings, HGS becomes significant, addressing diagnostic barriers and potentially revolutionizing diabetes management. However, limitations include a small sample size of 59 students and restrictions to specific demographics, emphasizing the need for future studies in diverse populations to validate HGS’s efficacy in real-world, resource-constrained settings.

## Introduction

Diabetes mellitus has emerged as one of the most pervasive health issues worldwide, driven by its global prevalence (1). Characterized by elevated blood sugar levels resulting from genetic factors, acquired deficiency, or insulin malfunction, diabetes significantly burdens healthcare systems. The number of diagnosed individuals with diabetes has been rapidly increasing, with a rise of 314 million cases from 1980 to 2014, reaching 415 million in 2014 (2). This alarming trend is projected to continue, with an estimated 625 million adults expected to be affected by diabetes by 2045, predominantly in low and middle-income countries, including several African nations (3).

Uncontrolled diabetes impairs patients’ quality of life and imposes substantial healthcare costs on countries (2). Data from the National Health and Nutritional Examination Survey (NHANES) reveal that the average lifetime medical costs for individuals with diabetes amount to as much as $85,200, with a significant portion dedicated to managing complications (4). Predictions highlight Africa as the region with the highest projected increase in the burden of diabetes and associated complications, despite contributing the least to global annual healthcare expenses for diabetes care. In 2017, the International Diabetes Federation (IDF) estimated the total health expenditure due to diabetes at $3.3 billion. In Nigeria alone, the national annual direct costs of diabetes were estimated to range from $1.071 billion to $1.639 billion (5).

Diabetes and its associated complications are responsible for more than 3 million deaths worldwide each year. In the United States of America, diabetes is the seventh leading cause of death, contributing to 69,091 deaths and playing a role in an additional 234,051 deaths (6). In Africa, more than 298,160 deaths, accounting for 6% of all mortality, were attributed to diabetes in 2017, with the highest proportion of all-cause mortality due to diabetes occurring in the age group of 30-39. Additionally, 77.0% of all deaths attributable to diabetes occurred in individuals under 60 years old, marking the highest proportion worldwide (5).

Diabetes is associated with numerous life-threatening complications and adverse health outcomes that develop gradually. These include neuropathy, skin complications, eye complications, diabetic ketoacidosis, gastroparesis, and macrovascular diseases (7).

In resource-constrained settings, access to primary or preventative healthcare is hindered by various barriers, such as a shortage of trained physicians and prohibitively high transportation costs. As a result, individuals often receive treatment once their conditions have reached a dangerously severe stage. Subsequently, many developing regions have implemented Community Health Worker (CHW) programs to bridge the gap between communities and healthcare providers. CHWs, typically volunteers, are trained to provide pre-primary healthcare and basic health information to rural communities lacking access to trained healthcare professionals. These dedicated individuals serve as trusted community leaders, mentors, and educators, working towards improving the health of their communities (8).

While CHWs have made significant strides in improving community health, their ability to effectively screen and diagnose diseases is limited by the need for more contextually appropriate tools and devices. Biomedical devices must therefore be affordable, ruggedized, and user-friendly. However, only some existing devices meet these criteria. For instance, current blood glucometers used to diagnose diabetes are expensive, requiring blood samples that pose health hazards. These devices often remain unused due to financial constraints faced by patients and healthcare professionals’ inability to afford test strip upkeep (4).

An alternative screening tool that shows promise in resource-constrained settings is handgrip strength, a simple measure of muscle strength that correlates well with other strength measures, such as quadriceps strength (8). Handgrip strength has been associated with metabolic syndrome, type 2 diabetes mellitus, and overall mortality (9); (10); (11). It indicates overall strength and physical activity level, as it measures the force produced by the muscles controlling the hand using a hand dynamometer (12).

Although the underlying mechanism is not completely understood, studies have explored the role of muscle resistance exercises in glucose metabolism and reported that such activities improve muscle function and glucose deposition, favouring insulin-mediated glucose uptake in skeletal muscle (13). Considering its relevance to various diseases like diabetes, malnutrition, and functional disability, handgrip strength testing with affordable and durable hand dynamometers has gained prominence (4).

Handgrip strength emerges as a promising and easy-to-measure health indicator suitable for screening diabetes in resource-constrained settings. Unlike expensive and hazardous diagnostic tests, handgrip strength testing avoids health risks associated with chemicals or bodily fluids. It offers a preventive and cost-effective approach to managing diabetes mellitus, particularly in developing countries like Nigeria, where access to healthcare is limited (4). By utilizing handgrip strength as a screening tool, barriers to diagnosis, such as high costs and limited access to healthcare professionals, can be overcome, facilitating early identification and intervention in high-risk populations.

Therefore, this article aims to investigate the relationship between handgrip strength and blood glucose regulation in non-diabetic young adults. By establishing this connection, the study intends to provide valuable insights into the potential of handgrip strength as a preventive and affordable approach to managing diabetes, ultimately reducing the economic implications of the disease, particularly in resource-constrained settings.

## Materials and methods

### Participants

One hundred students from the University of Ilorin, Nigeria, were initially recruited for this study. The recruitment process started on the 29^th^ of March to the 5^th^ of July, 2023, and was conducted through advertisements on social platforms, and participants were selected on a “first come” basis. All samples were collected and procedures carried out on the 15^th^ and 16^th^ of July, 2023. Due to incomplete data, information from only fifty-nine of the recruited students were used for the final computation and analysis of results.

### Inclusion criteria

The data collected for this study included currently enrolled students aged 18-30 years who exhibited normoglycemia with fasting blood glucose levels ranging from 70-100 mg/dL. Participants were also required to have no significant health conditions or physical impairments that could affect their grip strengths or fasting blood glucose levels.

### Exclusion criteria

Students with missing information, a history of elevated blood glucose or a diagnosis of diabetes, and those who were unwilling or unable to undergo handgrip strength measurements as part of the study protocol were excluded from the analysis.

### Ethical considerations

Self-reported questionnaires were used to assess baseline socio-demographic and lifestyle characteristics, existing medical conditions and use of medications. Ethical approval was collected from the Research Ethics Committee of the University of Ilorin Teaching Hospital, Ilorin Kwara state, with the reference number: UITH/CAT/189/VOL.21B /486.The research protocol was reviewed and approved by the relevant institutional review board. Informed and signed consent was obtained from all participants before their inclusion in the study.”

### Dependent variables

In this study, the dependent variables were glycaemic control and insulin resistance among non-diabetic students. As indicators, glycaemic control was assessed using HbA1c, fasting blood glucose, and 2-hour postprandial blood glucose. HbA1c is a marker for hyperglycaemia and provides information about blood plasma glucose levels over 2–3 months. An HbA1c above 7% and a 2-hour postprandial plasma glucose greater than 140 mg/dL were considered indicators of poor glycaemic control (14).

### Glycaemic control

HbA1c, also known as glycated haemoglobin, is formed when haemoglobin is exposed to plasma glucose through non-enzymatic pathways. It serves as a marker for hyperglycaemia and monitors blood plasma glucose levels over a prolonged period. Several factors, such as a high-fat diet, smoking (15), and body fat (16), can influence HbA1c levels. Two-hour postprandial blood glucose, measured two hours after a meal, is an essential indicator of postprandial plasma glucose levels, which play a significant role in overall glycaemic control.

### Insulin resistance

A fasting serum insulin above 10 µIU/mL was diagnostic of insulin resistance (17). In addition to serum insulin, fasting venous blood samples were collected to measure plasma glucose, C-peptide, and glycated haemoglobin levels. Plasma glucose was measured using a modified hexokinase enzymatic method, serum insulin was measured by radioimmunoassay, and glycated haemoglobin was measured using high-performance liquid chromatography (18).

### Independent variables

#### Handgrip strength

Handgrip strength (HGS) was measured as an indicator of muscle strength and functional capacity in daily activities. It was assessed using a dynamometer and is associated with various chronic diseases (19), cognitive decline (20), length of hospital stays, and mortality. Before measuring handgrip strength, participants were given instructions and a warm-up for their hands and fingers. The measurements were taken while participants stood with their feet hip-width apart and arms straight, slightly away from the body. Each hand was tested thrice, with a rest period between trials (21).

#### Relative handgrip strength

Relative HGS was calculated by dividing absolute HGS (kg) by BMI (reported as kg/BMI). This measure was used to adjust for the relationship between mass and force, considering both muscle quality and the combined effect of fat mass and muscle mass (22).

#### Height and body weight

Height and body weight were measured using standardized procedures (23). Participants’ standing height was measured with a stadiometer, and body weight was measured using a digital scale. Body mass index (BMI) was calculated by dividing the body weight (kilograms) by the square of height (meters) (kg/m²).

#### Waist/hip ratio

Waist circumference and hip circumference were measured using tape. Waist circumference was measured between the narrowest point between the ribs and hips, while hip circumference was measured at the point where the buttocks extended the most (24). Two consecutive recordings were made for each site.

#### Co-variates

The covariates in this study included sociodemographic characteristics, lifestyle factors, and self-reported family history of diseases. Sociodemographic characteristics covered age (years, continuous), gender (male/female), country, and ethnicity. Lifestyle factors included self-reported, exercise, drinking and smoking status.

#### Statistical analysis

A cross-sectional analysis was conducted and recorded as means (standard deviations) for continuous variables and frequencies for categorical variables. Differences between groups were assessed using ANOVAs or chi-square tests for continuous or categorical variables. Multiple linear regression models were used to examine the association between glucose regulation and grip strength. IBM SPSS Statistics Version 25.0 (IBM Corp., Armonk, New York, USA) was used for all statistical analyses.

## Results

Overall, data of 59 subjects (male 30 = 50.8 % and female 29 = 49.2 %) were used for this study with a mean age of 18 to 21 years. Dominant HGS ranged from 11.5 - 29.8 kg with interquartile range (IQR) of 18.4 – 25.1 kg (6.7 kg) in females and from 15.0 – 33.2 kg with IQR of 21.6 – 27.4 kg (5.8 kg) in males (Table 1).

**Table 1:** Sample Clinical Characteristics and biomarkers showing Mean, Quartiles, Interquartile Range & Standard deviation of Continuous variables and Percent (%) of Categorical variables (N = 59)

|  | Mean | Median | Min | Max | 25% | 75% | Interquartile Range (%) | Std Dev |
| --- | --- | --- | --- | --- | --- | --- | --- | --- |
| HGS Right Hand (kg) | 22.95 | 22.70 | 11.50 | 33.2 | 20.1 | 26.2 | 6.1 | 4.534 |
| HGS Left Hand (kg) | 21.46 | 21.20 | 13.80 | 31.80 | 18.2 | 24.9 | 6.7 | 4.543 |
| Absolute HGS (kg) | 46.22 | 45.60 | 28.40 | 67.80 | 41.4 | 52.4 | 11.0 | 8.706 |
| Relative HGS (m²) | 2.13 | 2.12 | 0.98 | 3.81 | 1.82 | 2.39 | 0.57 | 0.520 |
| BMI (Kg/m²) | 22.32 | 22.00 | 15.40 | 38.6 | 19.4 | 23.8 | 4.4 | 4.304 |
| Waist/Hip Ratio | 0.79 | 0.80 | 0.70 | 1.1 | 0.7 | 0.80 | 0.1 | 0.078 |
| Pulse Pressure (mmHg) | 49.34 | 49.00 | 30.0 | 68 | 43.0 | 57.0 | 14.0 | 9.278 |
| Fasting Blood Glucose (mmol/L) | 4.86 | 4.80 | 3.60 | 6.4 | 4.5 | 5.3 | 0.8 | 0.560 |
| HBA1c (%) | 3.73 | 3.42 | 0.51 | 8.58 | 2.87 | 4.59 | 1.72 | 1.396 |
| 2-hour Postprandial Glucose (mmol/L) | 5.20 | 5.20 | 3.90 | 7.6 | 4.5 | 5.7 | 1.2 | 0.779 |
| Serum Insulin (µIU/L) | 18.86 | 14.09 | 5.88 | 158.55 | 10.58 | 19.59 | 9.01 | 20.522 |
|  | N | (%) |  |  |  |  |  |  |
| Gender |  |  |  |  |  |  |  |  |
| Male | 30 | 50.8 |  |  |  |  |  |  |
| Female | 29 | 49.2 |  |  |  |  |  |  |
| Age |  |  |  |  |  |  |  |  |
| 18-21 | 19 | 32.2 |  |  |  |  |  |  |
| 22-25 | 38 | 64.4 |  |  |  |  |  |  |
| 26-30 | 2 | 3.4 |  |  |  |  |  |  |
| Smokes |  |  |  |  |  |  |  |  |
| Yes | 0 | 0.0 |  |  |  |  |  |  |
| No | 59 | 100.0 |  |  |  |  |  |  |
| Alcohol Intake |  |  |  |  |  |  |  |  |
| Yes | 6 | 10.2 |  |  |  |  |  |  |
| No | 53 | 89.8 |  |  |  |  |  |  |
| Does Exercise |  |  |  |  |  |  |  |  |
| Yes | 29 | 49.2 |  |  |  |  |  |  |
| No | 30 | 50.8 |  |  |  |  |  |  |
| Hand Dominance |  |  |  |  |  |  |  |  |
| Right | 51 | 86.4 |  |  |  |  |  |  |
| Left | 8 | 13.6 |  |  |  |  |  |  |
| BMI denotes body mass index |  |  |  |  |  |  |  |  |
| HBA1c denotes glycated haemoglobin |  |  |  |  |  |  |  |  |
| HGS denotes Handgrip strength |  |  |  |  |  |  |  |  |

In this study HGS < 18kg was defined as low while HGS >18kg was defined as normal, fasting blood sugar between 3.9-5.9mmom/l and 2HPG < 7.8 mmol/l was defined as normal range of blood glucose levels. HGS values recorded from study subjects were within normal range with a mean of 21.07kg and 18.70kg for dominant and non-dominant hands respectively

The dominant HGS in females (mean = 21.4 ± 4.53) was significantly reduced (p = 0.005) when compared to males (mean = 24.6 ± 4.06). (Table 2). Non-Dominant HGS ranged from 13.8 - 25.8 kg with interquartile range (IQR) of 15.7 – 21.2 kg (5.5 kg) in females and from 14.7 – 31.8 kg with IQR of 20.5 – 26.1 kg (5.6 kg) in males. The non-dominant HGS in females (mean = 18.9 ± 3.61) was significantly reduced (p = 0.001) when compared to males (mean = 24.0 ± 4.09). (Table 2)

**Table 2:** Clinical Characteristics and biomarkers by sex.

|  | **Males** |  | **Females** |  |  |
| --- | --- | --- | --- | --- | --- |
|  | Mean | Std Dev | Mean | Std Dev | p value |
| **HGS Right Hand (kg)** | 24.62 | 4.06 | 21.37 | 4.53 | 0.008 |
| **HGS Left Hand (kg)** | 24.01 | 4.09 | 18.97 | 3.61 | 0.000 |
| **Absolute HGS (kg)** | 4975 | 8.27 | 42.87 | 7.99 | 0.003 |
| **Relative HGS (m<sup>2</sup>)** | 2.37 | 0.61 | 1.93 | 0.40 | 0.003 |
| **BMI (Kg/m<sup>2</sup>)** | 21.59 | 3.47 | 22.85 | 5.13 | 0.357 |
| **Waist/Hip Ratio** | 0.82 | 0.06 | 0.77 | 0.09 | 0.020 |
| **Pulse Pressure (mmHg)** | 53.65 | 8.02 | 44.55 | 8.14 | 0.000 |
| **Fasting Blood Glucose (mmol/L)** | 4.69 | 0.64 | 5.01 | 0.44 | 0.042 |
| **HBA1c (%)** | 3.81 | 1.24 | 3.61 | 1.56 | 0.519 |
| **2-hour Postprandial Glucose (mmol/L)** | 5.15 | 0.75 | 5.22 | 0.83 | 0.818 |
| **Serum Insulin (μIU/L)** | 17.40 | 8.76 | 15.65 | 9.01 | 0.449 |
|  | **N = 30** | **%** | **N = 29** | **%** |  |
| **Age** |  |  |  |  |  |
| **18-21** | 6 | 20.0 | 13 | 44.83 |  |
| **22-25** | 23 | 77.0 | 15 | 51.72 |  |
| **26-30** | 1 | 3.0 | 1 | 3.45 |  |
| **Smokes** |  |  |  |  |  |
| **Yes** | 0 | 0.0 | 0 | 0.00 |  |
| **No** | 30 | 100.0 | 29 | 100.00 |  |
| **Alcohol Intake** |  |  |  |  |  |
| **Yes** | 1 | 3.3 | 5 | 17.24 |  |
| **No** | 28 | 93.4 | 24 | 82.76 |  |
| **Missing** | 1 | 3.3 | 0 | 0.00 |  |
| **Does Sports** |  |  |  |  |  |
| **Yes** | 19 | 63.3 | 10 | 34.48 |  |
| **No** | 11 | 36.7 | 19 | 65.52 |  |
| **Hand Dominance** |  |  |  |  |  |
| **Right** | 27 | 90.0 | 25 | 86.21 |  |
| **Left** | 3 | 10.0 | 4 | 13.79 |  |

In both sexes, there was significant difference (female p = 0.03 & male p = 0.04) (Fig 1) in HGS between both hands suggesting that hand dominance could be a relevant factor in this study. As such, the results of dominant and non-dominant HGS were also considered independently (Table 3).

**Figure 1:**
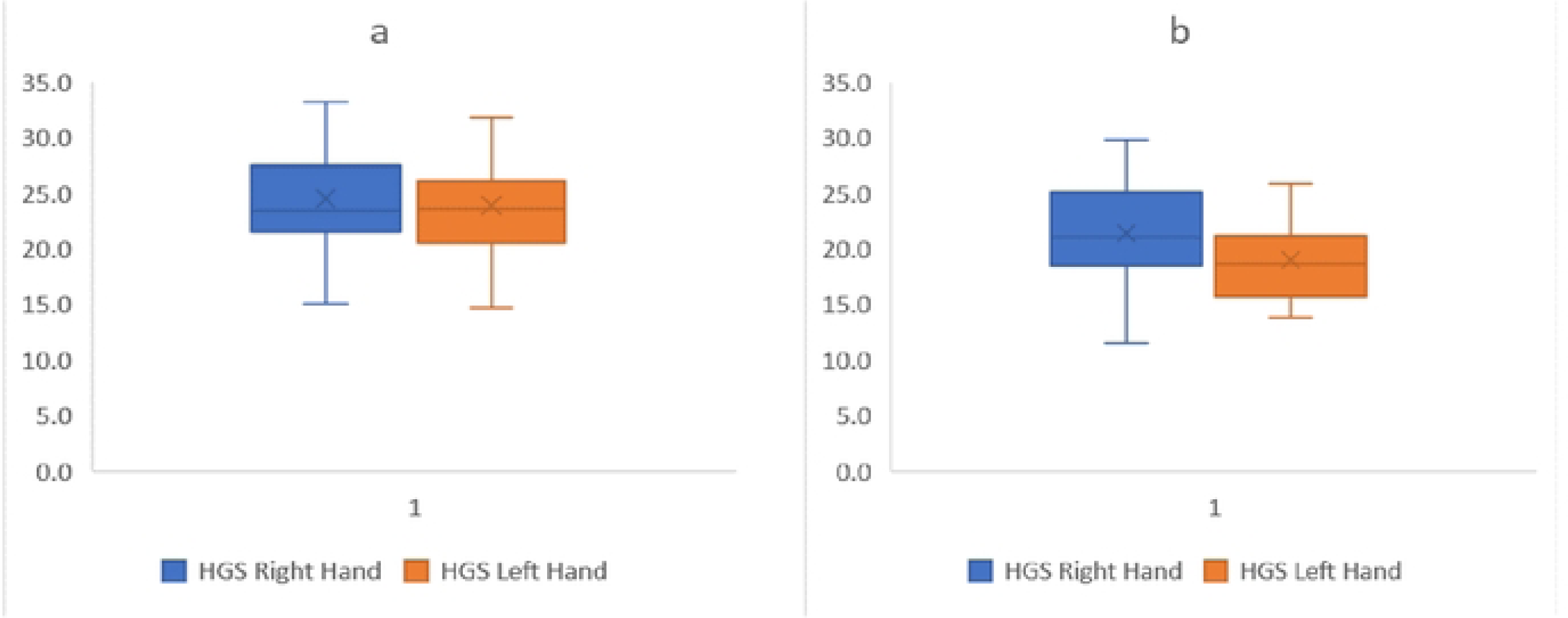
Boxplots comparing handgrip strength in both dominant and non-dominant hands in both male (a) and female (b) subjects. Mean HGS in males was 24.47 ± 4.05 and 23.86 ± 4.07 in both right and left hands, respectively; and in females was 21.37 ± 4.53 and 18.97 ± 3.61 in both right and left hands, respectively. The difference was statistically significant at p-value 0.03 and 0.04 between hands in females and males, respectively. HGS – handgrip strength

**Table 3:** Results of Multiple linear regression of handgrip strength (dominant and non-dominant) on blood glucose regulation biomarkers.

|  | Male Handgrip Strength |  |  |  | Female Handgrip strength |  |  |  |
| --- | --- | --- | --- | --- | --- | --- | --- | --- |
|  | Dominant |  | Non-dominant |  | Dominant |  | Non-dominant |  |
|  | Estimate (SE) | p | Estimate (SE) | p | Estimate (SE) | P | Estimate (SE) | P |
| Fasting Blood Glucose | 0.3758 (0.59) | **0.04** | 0.3941 (0.61) | 0.07 | 0.3218 (0.42) | 0.09 | 0.2330 (3.58) | 0.22 |
| 2-hour Post Prandial | 0.1117 (0.76) | 0.55 | 0.1049 (0.76) | 0.57 | 0.3407 (0.78) | 0.07 | 0.3887 (3.39) | **0.04** |
| HBA1c | 0.0184 (1.26) | 0.92 | 0.1277 (1.25) | 0.57 | 0.0587(1.58) | 0.76 | 0.2090 (3.60) | 0.28 |
| Serum Insulin | 0.2303 (8.82) | 0.22 | 0.2226 (8.83) | 0.22 | 0.0846 (9.14) | 0.66 | 0.0678 (28.48) | 0.73 |

Multiple regression analysis was employed to examine the relationships between handgrip strength and the blood glucose regulatory markers, specifically fasting blood glucose, 2 hours postprandial glucose, HBA1c and serum insulin levels. Four different models were tested to account for potential confounders: Model 1 (no adjustments), Model 2 (adjusted for WHR), Model 3 (adjusted for BMI), and Model 4 (adjusted for both WHR and BMI) (Tables 4, 5 and 6).

**Table 4:** Results of Multiple regression of absolute handgrip strength and relative handgrip strength on blood glucose regulation biomarkers.

|  | Absolute Handgrip Strength |  |  |  | Relative Handgrip strength |  |  |  |
| --- | --- | --- | --- | --- | --- | --- | --- | --- |
|  | Male |  | Female |  | Male |  | Female |  |
|  | Estimate (SE) | p | Estimate (SE) | p | Estimate (SE) | P | Estimate (SE) | P |
| Fasting Blood Glucose | 0.322 (0.62) | 0.09 | 0.319 (0.42) | 0.08 | 0.139 (0.64) | 0.46 | 0.08 (0.45) | 0.68 |
| 2-hour Post Prandial | 0.067 (0.76) | 0.72 | 0.396 (0.77) | **0.03** | 0.287 (0.73) | 0.12 | 0.284 (0.80) | 0.14 |
| HbA1c | 0.088 (1.25) | 0.64 | 0.085 (1.58) | 0.66 | 0.335 (1.19) | 0.07 | 0.303 (1.51) | 0.11 |
| Serum Insulin | 0.232 (8.67) | 0.21 | 0.102 (9.12) | 0.59 | 0.227 (8.68) | 0.22 | 0.079 (9.14) | 0.68 |

SE denotes standard error. HbA1c denotes glycated haemoglobin

**Table 5:** Adjusted Relationships of Handgrip Strength With blood glucose regulatory markers (males n=29)

|  | Fasting blood glucose |  | 2-hour post prandial |  | HbA1c |  | Serum Insulin |  |
| --- | --- | --- | --- | --- | --- | --- | --- | --- |
| Absolute HGS | Estimate (SE) | P | Estimate | P | Estimate | P | Estimate | P |
| Model 1<sup>a</sup> | 0.3755 (0.59) | **0.04** | 0.1133 (0.76) | 0.55 | 0.0560 (1.26) | 0.77 | 0.2318 (8.82) | 0.22 |
| Model 2<sup>b</sup> | 0.4543 (0.58) | **0.04** | 0.2511 (0.75) | 0.42 | 0.2165 (1.26) | 0.52 | 0.3694 (8.58) | 0.14 |
| Model 3<sup>c</sup> | 0.5311 (0.56) | **0.01** | 0.4231 (0.70) | 0.07 | 0.3239 (1.22) | 0.22 | 0.2435 (8.95) | 0.44 |
| Model 4<sup>d</sup> | 0.5465 (0.56) | **0.03** | 0.4311 (0.71) | 0.14 | 0.3392 (1.23) | 0.36 | 0.3707 (8.74) | 0.27 |
| Relative HGS |  |  |  |  |  |  |  |  |
| Model 1<sup>a</sup> | 0.0436 (0.64) | 0.82 | 0.2223 (0.74) | 0.24 | 0.2947 (1.21) | 0.11 | 0.2408 (8.79) | 0.20 |
| Model 2<sup>b</sup> | 0.2408 (0.64) | 0.45 | 0.2769 (0.75) | 0.34 | 0.3248 (1.22) | 0.22 | 0.3406 (8.68) | 0.19 |

**Note: Boldface indicates statistical significance (p<0.05).**
SE denotes standard error. HbA1c denotes glycated haemoglobin
<sup>a</sup> Multiple Linear regression analysis
<sup>b</sup> Adjusted for Waist hip ratio (WHR)
<sup>c</sup> Adjusted for Body mass index (BMI)
<sup>d</sup> Adjusted for WHR and BMI

**Table 6.** Adjusted Relationships of Handgrip Strength With blood glucose regulatory markers (females n=30)

|  | Fasting blood glucose |  | 2-hour post prandial |  | HBA1c |  | Serum Insulin |  |
| --- | --- | --- | --- | --- | --- | --- | --- | --- |
| Absolute HGS | Estimate (SE) | P | Estimate | P | Estimate | P | Estimate | P |
| Model 1<sup>a</sup> | 0.319 (0.42) | 0.08 | 0.396 (0.77) | **0.03** | 0.085 (1.58) | 0.66 | 0.102 (9.12) | 0.59 |
| Model 2<sup>b</sup> | 0.3683 (0.42) | 0.15 | 0.4243 (0.77) | 0.08 | 0.1553 (1.59) | 0.73 | 0.1671 (9.21) | 0.69 |
| Model 3<sup>c</sup> | 0.4641 (0.40) | **0.04** | 0.4336 (0.77) | 0.07 | 0.4801 (1.42) | **0.03** | 0.1037 (9.29) | 0.87 |
| Model 4<sup>d</sup> | 0.4872 (0.41) | 0.08 | 0.4541 (0.78) | 0.12 | 0.4866 (1.44) | 0.08 | 0.1672 (9.39) | 0.87 |
| Relative HGS |  |  |  |  |  |  |  |  |
| Model 1<sup>a</sup> | 0.0800 (0.45) | 0.68 | 0.2840 (0.80) | 0.14 | 0.3030 (1.51) | 0.11 | 0.0790 (9.14) | 0.68 |
| Model 2<sup>b</sup> | 0.1626 (0.45) | 0.71 | 0.3291 (0.81) | 0.23 | 0.3176 (1.53) | 0.25 | 0.1567 (9.23) | 0.72 |

**Note: Boldface indicates statistical significance (p<0.05).**
SE denotes standard error. HBA1c denotes glycated haemoglobin
<sup>a</sup> Multiple Linear regression analysis
<sup>b</sup> Adjusted for Waist hip ratio (WHR)
<sup>c</sup> Adjusted for Body mass index (BMI)
<sup>d</sup> Adjusted for WHR and BMI

### Findings in males and females

While still maintaining normal ranges in the blood regulatory markers (FBS, 2-hour Postprandial and HBA1c) serum insulin levels were slightly elevated in both sexes (male 17.40±8.76 female 15.65±9.01).

### Fasting blood glucose

In males, a notable finding emerged as absolute handgrip strength was consistently linked to fasting blood glucose levels across all models (P<0.05), irrespective of adjustments made for WHR and BMI (Table 5). This association persisted, highlighting the robustness of the relationship. In contrast, among females, absolute HGS was only found to be associated to blood glucose levels following adjustments to BMI (Model 3) (Table 6).

### 2-hour post-prandial glucose

The investigation into the relationships between handgrip strength and 2-hour post-prandial glucose levels showed a significant (p<0.05) association in females. Notably, no significant relationships were observed in males, regardless of the adjustments made for potential confounding factors.

### HBA1c

For females, an interesting finding emerged in Model 3, where adjustments were made for BMI. In this scenario, a significant positive relationship was observed between absolute HGS and HBA1c levels. This indicates that higher HGS may be associated with higher HBA1c levels when considering BMI as a confounding factor. However, it is important to note that this association was not observed in males or in other models.

### Serum insulin

Irrespective of gender, our analyses found no significant associations between HGS and serum insulin levels across all models tested. These results suggest that HGS may not be a strong predictor of serum insulin levels in our study cohort.

## Discussion

The primary findings of our study reveal nuanced relationships between handgrip strength and various blood markers related to diabetes, including fasting blood glucose, 2-hour post-prandial glucose, HBA1c, and serum insulin levels. The results are particularly compelling in the context of elevated serum insulin levels observed in both male and female participants, despite other blood regulatory markers remaining within normal ranges.

Our study presents intriguing findings regarding the correlation between handgrip strength and fasting blood glucose levels in males and females. In males, the association remained robust and statistically significant across all models, even after adjusting for waist-hip ratio and body mass index. This suggests that handgrip strength could be a reliable marker for glucose metabolism in this demographic. Contrastingly, in females, the relationship became evident only after adjusting for BMI, indicating that body composition plays a significant role in mediating this relationship.

Several mechanisms could explain these associations. For males, the findings align with previous research emphasizing the role of enhanced muscle metabolism and higher testosterone levels in insulin sensitivity (25); (26). Other studies have further elucidated the role of testosterone in promoting muscle glucose uptake and improving muscle function, thereby reinforcing its importance in glucose metabolism (27). Muscles are a significant site for glucose uptake, and efficient neuromuscular junctions may facilitate more effective muscle contractions, thereby demanding more glucose (28) This suggests that a more substantial handgrip indicates better neuromuscular junction efficiency, which could affect metabolic processes like glucose regulation.

For females, the role of body composition is more complex. Our findings imply that the relationship between HGS and glucose metabolism might be confounded by factors like body fat percentage, which is generally higher in females (29). This corresponds to studies that have indicated that increased adiposity can lead to insulin resistance and consequently disrupt glucose homeostasis (30). The fact that the relationship became significant only after BMI adjustments suggests that body composition, particularly fat mass, may be a critical mediator of this relationship in females. Indeed, adipose tissue is not just an energy storage organ but also an active endocrine organ that releases various factors, including adipokines, which can affect insulin sensitivity and glucose metabolism (31). These adipokines have been implicated in the pathogenesis of insulin resistance, particularly in females, where the balance between different adipokines can be more easily perturbed. The interplay between muscle and fat tissue in females could be more complex, given the roles of adipokines and other hormones that influence insulin sensitivity (31). This likely reflects a more intricate physiological interaction that warrants further investigation.

Our study found no correlation between handgrip strength and serum insulin levels in line with Niemann et al. findings (32), while another study led by Lazarus et al. in 1997 (33) reported a modest correlation between these variables. Our findings highlight that the relationship between muscle strength and insulin levels can vary based on specific populations or experimental methodologies. Since skeletal muscles are primary sites for insulin-stimulated glucose uptake, it is logical to assume that stronger muscles could be more efficient in glucose uptake, influencing insulin levels. However, our results suggest that different physiological mechanisms might modulate muscle strength and insulin functions. Skeletal muscle mass balance is a function of protein synthesis and breakdown. Factors like fasting, trauma, or specific disease states can accelerate muscle protein breakdown, as shown in various studies. Insulin, a pivotal hormone, regulates muscle protein breakdown by affecting the transcription of crucial proteins, as evidenced by studies on FOXO transcription factors (34).

Thus, while there are clear links between muscle strength and the efficiency of skeletal muscle in using glucose, and muscle mass and insulin, it seems that muscle strength might not directly affect insulin secretion or function, which involves a more complex interplay of factors.

We identified a notable association between handgrip strength and 2-hour post-prandial glucose levels, but this was evident only in females and not males. A similar study by Huang in 2023 emphasized that the effect of handgrip strength on Type 2 Diabetes Mellitus (T2DM) could be influenced by factors such as BMI and gender (35). This gender divergence in results underlines the need to consider gender-specific physiological pathways when using handgrip strength as a diabetes screening tool.

For females, the significant correlation may be attributed to the role of estrogen, which is known to modulate muscle function and insulin sensitivity. Studies by Chidi-Ogbolu & Baar (36), and Camporez et al., (37), support this assertion, indicating estrogen’s potential to enhance insulin-stimulated glucose uptake in muscles (38).

On the other hand, the relationship in males is more intricate due to testosterone’s fluctuating effects on insulin sensitivity. While testosterone’s influence on muscle strength is well-documented, its impact on insulin sensitivity can vary based on age and general health. This observation aligns with findings by Dhindsa et al., 2016 (39), complicating the establishment of a direct link between handgrip strength and post-prandial glucose levels in males.

Our study’s salient observation is the link between handgrip strength and HbA1c levels, especially when considering BMI. This association sheds light on the intricate interplay of hormonal and metabolic factors in the human body. Similarly, Mainous III et al., 2015, highlighted that handgrip strength negatively correlated with HbA1c levels (40) strengthening the credibility of HbA1c as a marker for prolonged glucose control.

The association between muscle, fat tissue, and glucose regulation is persistent, indicating the importance of handgrip strength as a potential indirect indicator of long-term glycemic control in areas with limited resources. This assertion is consistent with the findings of Jang et al., 2020, who explored the relationship between relative handgrip strength and prediabetes based on HbA1c levels and emphasized the significance of sex differences (38).

The influence of hormones like testosterone and estrogen on body fat distribution and muscle metabolism plays a pivotal role in understanding this association. While testosterone generally promotes abdominal fat storage and muscle growth, estrogen affects fat storage in the hips and thighs, alongside its distinct role in muscle metabolism. These hormonal influences underline the complexities of long-term glucose regulation, as manifested by HbA1c levels.

Muscle and fat tissues have unique metabolic contributions. While muscle tissue, being metabolically active, is crucial for glucose uptake, fat tissue releases adipokines that might alter insulin sensitivity. Together with hormones such as insulin and thyroid hormone, these factors intricately shape the observed relationship between handgrip strength and HbA1c levels.

## Conclusion

### Summary of Key Findings

Our study has introduced an innovative perspective on handgrip strength as a screening tool for managing blood glucose regulation disorders. This pioneering approach aligns with existing literature, establishing a solid association between HGS and blood glucose regulatory markers. Importantly, our research reveals the potential of HGS assessments as practical and cost-effective means to identify individuals at risk of blood glucose irregularities, including diabetes. By incorporating HGS assessments into healthcare protocols, timely interventions, including exercise-based programs, can be initiated, offering a personalized approach to blood glucose management.

### Implications for Resource-Constrained Settings

In resource-constrained settings, the utility of HGS as an accessible, non-invasive, and cost-effective screening tool becomes particularly significant. This approach addresses the barriers to diagnosis, enabling Community Health Workers to conduct HGS tests using inexpensive and readily available hand dynamometers. Our study has the potential to revolutionize diabetes management in developing countries, providing a viable solution to overcome diagnostic limitations and reduce the economic burden associated with the disease. However, it is vital to acknowledge the need for tailored interventions, considering the complexity of hormonal and metabolic factors in diverse populations.

### Study Limitations

While our findings are promising, acknowledging the study’s limitations is crucial. The sample size of 59 students may not fully represent the broader population, potentially affecting the study’s statistical power. A more extensive and diverse sample would strengthen the results and minimize the risk of overlooking potential relationships (type II errors). Furthermore, the study’s restriction to students may limit its applicability to various age groups, occupations, and demographic factors. Additionally, lifestyle, nutritional status, and other determinants may differ significantly from the student cohort in resource-constrained settings. Therefore, caution is necessary when generalizing these findings to broader contexts. Future studies with more diverse and larger participant groups must validate these findings and ensure the screening tool’s efficacy in real-world, resource-constrained settings.

## Data Availability

All relevant data are within the manuscript and its Supporting Information Files

## Acknowledgements

We thank all the participants and research staffs of the Physiology Laboratory of the University of Ilorin, Ilorin, Nigeria, for their helpful cooperation in this study.

## Supporting information

**S1 Fig 1 Title**

Boxplots comparing handgrip strength in both dominant and non-dominant hands in both male and female subjects

**Si Fig 1 Legend**

### Ethical approval

Ethical approval was collected from the Research Ethics Committee of the **University of Ilorin Teaching Hospital,** Ilorin Kwara state, with the reference number: UITH/CAT/189/VOL.21^B^/486. An informed consent was also obtained from the research subjects.

